# A Prospective Natural History Study Protocol for Clinical Trial Readiness in Synaptic Disorders

**DOI:** 10.64898/2026.01.30.26344887

**Authors:** Jillian L. McKee, Sarah M. Ruggiero, Kristin Cunningham, JoeyLynn Coyne, Ian McSalley, Michael C. Kaufman, Bintou Bane, Torrey Chisari, Jonathan Toib, Carlyn Glatts, Sarah Tefft, Julie M. Orlando, Viveknarayanan Padmanabhan, Alexander K. Gonzalez, Alicia Harrison, Charlene Woo, Stephanie A. Zbikowski, Rency Dhaduk, Johanna Mercurio, Macie McCarthy, Jan H. Magielski, Zachary Grinspan, Megan Abbott, Juliet Knowles, Hsiao-Tuan Chao, Katherine Xiong, Elizabeth Berry-Kravis, Sepideh Tabarestani, J. Michael Graglia, Kathryn Helde, Virginie McNamar, Charlene Son Rigby, James Goss, Scott Demarest, Andrea Miele, Benjamin Prosser, Michael J. Boland, Samuel R. Pierce, Ingo Helbig

## Abstract

**Objective:** *STXBP1-*Related Disorders (*STXBP1*-RD) and *SYNGAP1*-Related Disorder (*SYNGAP1*-RD) are two common genetic synaptopathies, leading to epilepsy, developmental delay, and intellectual disability. Both *STXBP1*-RD and *SYNGAP1*-RD are potential targets for disease-modifying therapies, but there is limited information in the literature describing the natural history of either disorder, which impedes outcome selection for future clinical trials. The objective of this study is to develop a framework to better define and outline the clinical spectrum and longitudinal trajectories of *STXBP1*-RD and *SYNGAP1*-RD natural history, including development, behavior, seizure histories, and electrophysiology.

**Methods:** Here, we describe a protocol, regulatory structure, and supportive preliminary data for multi-center, prospective natural history studies of *STXBP1*-RD (STARR) and *SYNGAP1*-RD (ProMMiS). The protocols incorporate gold-standard clinician-assessed outcome measures including the Bayley Scales of Infant and Toddler Development 4^th^ edition, Gross Motor Function Measure-66, and fine motor domains of the Peabody Developmental Motor Scales 3^rd^ Edition, parent reported outcome measures (PROMs), epilepsy histories, and biomarker exploration. To date, the study has enrolled 164 individuals with *STXBP1*-RD and 159 with *SYNGAP1*-RD, with ongoing longitudinal assessments every 6 months in a subset of approximately 200 total individuals across both disorders.

**Results:** Our data support that existing developmental measures are feasible, informative, and show minimal floor or ceiling effects. Furthermore, we demonstrate that medical record-based seizure history reconstruction reveals unique epilepsy trajectories while minimizing burden to families. We observe disease-specific patterns of developmental performance and distinct longitudinal seizure dynamics, highlighting the need for data generation in a gene/disorder-specific manner for clinical trial readiness.

**Significance:** In summary, we present a feasible natural history protocol with prospective data for two complex neurodevelopmental disorders with natural histories that have previously been incompletely characterized, within a regulatory framework that will support the use of these data to expedite clinical trial development.

**Key Points:**

1. *STXBP1*-Related Disorder (*STXBP1*-RD) and *SYNGAP1*-Related Disorder (*SYNGAP1*-RD) are two common genetic causes of epilepsy, developmental delay, and intellectual disability.
2. *STXBP1*-RD and *SYNGAP1*-RD are potential targets for drug and gene therapy, but there is limited information in the literature describing the natural history of either disorder which impedes the development of possible therapeutics.
3. The current paper is a description of a prospective natural history study of *STXBP1*-RD and *SYNGAP1*-RD which will assess the clinical spectrum of each disorder through detailed developmental assessments, seizure histories, behavioral assessments, and electronic medical record reconstruction.
4. By studying *STXBP1*-RD and SYNGAP1-RD with both cross sectional and longitudinal assessments, we aim to improve clinical trial readiness so that potential treatments can be assessed expeditiously.

## Introduction

*STXBP1-* and *SYNGAP1*-related disorders (RD) are two of the most common genetic developmental and epileptic encephalopathies (DEE)[1, 2], and both have been established as prime targets for gene-targeted therapies given the severe burden of illness and haploinsufficiency as the disease mechanism[3]. Despite the relative prevalence of these two neurogenetic disorders, most of the scientific knowledge on clinical outcomes and seizure dynamics has been shaped by retrospective data[4-6]. Given the urgent need to develop high-quality, prospective natural history data for future clinical trials, we launched multi-site, prospective natural history studies for both *STXBP1-RD* and *SYNGAP1-RD*.

*STXBP1*-RD and *SYNGAP1*-RD are disorders of synaptic transmission resulting in a range of neurodevelopmental symptoms and epilepsy, with distinct phenotypic profiles. *STXBP1*-RD is predominantly characterized by global developmental delay and severe to profound intellectual disability, motor deficits driven by hypotonia and tremor, and neurobehavioral issues such as hyperactivity, aggression, and autism[4, 7, 8]. Approximately 80% of patients have some form of epilepsy, which most often starts in the neonatal or infantile periods, but has been seen to onset as late as the teenage years[9, 10]. Individuals with *STXBP1*-RD can variably present with multiple epilepsy *SYNGAP1*-RD is characterized by global developmental delay, intellectual disability, hypotonia, and generalized epilepsy often presenting as myoclonic-atonic epilepsy (EMAtS), and epilepsy with eyelid myoclonia (EEM), with onset typically in the toddler years[11-15]. *SYNGAP1*-RD also presents a complex neuropsychiatric profile that can include autism spectrum disorder, sleep difficulties, sensory processing disorders, and sometimes severe behavioral challenges[6, 11]. Current treatment options for *STXBP1*-RD and *SYNGAP1*-RD are limited and focused on symptom management. Antiseizure medications are the primary intervention for seizures, while drug resistance is common, resulting in frequent changes in medication over the lifespan[4, 8, 16, 17]. Unfortunately, there are currently no drugs available to directly or indirectly address the cognitive impairments and developmental delays associated with *STXBP1*-RD and *SYNGAP1*-RD.

Despite increased attention on *STXBP1*-RD and *SYNGAP1*-RD as targets for disease-modifying therapies, knowledge of the natural history of both disorders has been limited to retrospective phenotyping efforts and case reports[4-6]. Seizures are a common symptom among individuals with *STXBP1-*RD and *SYNGAP1*-RD, but disease concept models for both disorders have demonstrated that seizures present just one of many challenges for patients and caregivers, emphasizing the urgency to consider non-seizure outcomes as potential endpoints for future intervention studies[18]. A disease concept model for *STXBP1*-RD found that developmental delay, behavior, and seizures were major concerns for caregivers which affected the autonomy, socialization, and school performance of their children, while impacting their emotional state, need for support, and daily life[19]. Parents and caregivers of children with *SYNGAP1*-RD have reported that difficulty with motor skills, delays in language development, a high rate of autism spectrum disorder, and the presence of seizures were common features of the disorder[20].

Clinical trial readiness has become a prominent issue in the field. The development of precision therapies for rare neurodevelopmental disorders, such as anti-sense oligonucleotides (ASOs) and gene therapy, has rapidly expanded over the last several years, with numerous novel candidate therapies under development[3, 21, 22]. To evaluate the potential efficacy of these treatments, a detailed understanding of the natural histories of these disorders is required, an opinion which was also recently emphasized by guidance from the FDA[23]. Thus, it is imperative to fully describe a disease’s presentation at various timepoints and to see how the condition may change over time to fully inform inclusion criteria and outcome measure selection for future interventional trials[24].

Here we describe our prospective natural history protocols for *STXBP1*-RD (STARR) and *SYNGAP1*-RD (ProMMiS) and present initial data supportive of our study design. We aim to both develop a comprehensive study to characterize the natural history of *STXBP1*-RD and *SYNGAP1*- RD, while facilitating the creation of a natural history study framework that can be generalized to expedite clinical trial readiness in other DEEs more broadly.

## Methods

### Study Organization & Management

The structure of the natural history study for synaptic disorders is a prospective, multi-site, non-interventional study which will characterize the natural history in participants with *STXBP1*-RD and *SYNGAP1-*RD (**Figure 1**). Children’s Hospital of Philadelphia (CHOP) is the prime site and the Data Coordinating Center (DCC) for both studies. For the *STXBP1-*RD arm of the study (STARR), the sites include Baylor College of Medicine/Texas Children’s Hospital, Children’s Hospital Colorado, Stanford University Medical Center, Rush University Medical Center, Weill Cornell Medicine, and CHOP. For the *SYNGAP1-*RD arm (ProMMiS), study sites include Children’s Hospital Colorado, Stanford University Medical Center, and CHOP. All study sites received approval from their local Institutional Review Boards (IRBs), and the CHOP IRB holds agreements with each site principal investigator for regulatory oversight of respective activities.

**Figure 1.**
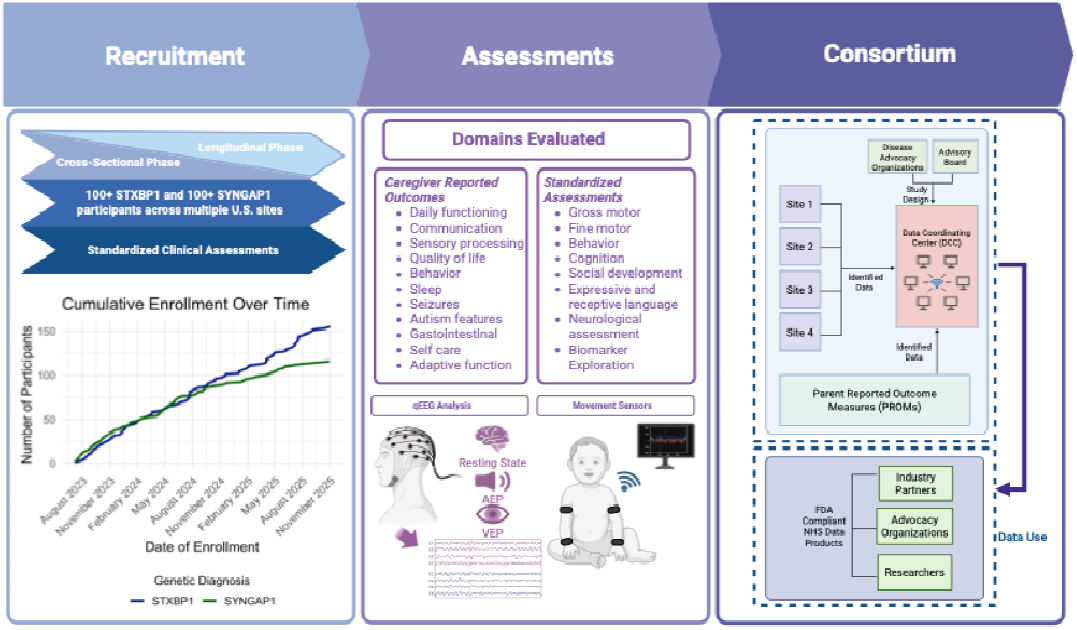
Protocol and Recruitment. Here we present a comprehensive and feasible protocol for evaluating the natural history of *STXBP1* (STARR) and *SYNGAP1* (ProMMiS) using existing outcome measures within the framework of a clinical visit. Recruitment is ongoing, with n=323 individuals evaluated to date. Study protocol includes standardized clinical assessments, parent-reported outcomes, quantitative EEG and movement sensors, and detailed seizure histories. Consortium structures have been established for both studies allowing data generation from multiple sites in an FDA-compliant format for collaboration with industry partners, advocacy organizations, and researchers.

### Study Design

The natural history study has two main aims—first, to cross-sectionally evaluate as many individuals as possible to fully capture the phenotypic breadth of the disorders and second, to follow a subset of 200 individuals longitudinally every 6 months to understand developmental trajectories and assess the stability of the measures over time. The natural history of each condition is captured alongside concurrent standard-of-care treatments, including but not limited to anti-seizure medications, physical and occupational therapy, speech-language therapy, behavioral therapy, dietary therapy, and surgical intervention.

### Recruitment

Subject recruitment in the ongoing natural history study follows various mechanisms. Eligible subjects at each site are referred by clinicians, who may include the study investigators. Recruitment is conducted in collaboration with patient advocacy groups, and includes outreach through postings on clinicaltrials.gov, social media, and sharing of study information through family organizations. Also, patients may be referred by outside clinicians to the study or through commercial laboratories who have performed diagnostic genetic testing. Once eligible subjects are identified, their medical record is reviewed for eligibility and the family or subject is then contacted. Parents or legal guardians of potential subjects are asked to sign a written consent before participating in the study. Due to the expected cognitive function of most children with *STXBP1*-RD and *SYNGAP1*-RD, a waiver for assent was obtained for the study. Adults who can provide consent or assent will sign the appropriate forms prior to participating. See **Table 1** for study inclusion and exclusion criteria.

**Table 1.** Inclusion/Exclusion Criteria.

| *Inclusion Criteria* |
| --- |
| • Male or female of any age<br>• Presence of a *STXBP1* or *SYNGAP1* causative genetic variant:<br>○ Variant must be classified as causative based on clinical and variant classification as determined by a genetic reviewer<br>○ Historical documentation of past result is sufficient<br>○ Confirmatory testing will be obtained, if necessary, at baseline and performed by a CLIA certified laboratory |
| *Exclusion Criteria* |
| • Presence of a confirmed mutation in a gene other than *STXBP1* or *SYNGAP1* that is known to contribute to a neurodevelopmental disability, including full gene deletions of *STXBP1* or *SYNGAP1* that include other genes beyond *STXBP1* or *SYNGAP1*<br>• Presence of a significant non-*SYNGAP1*-RD or non-*STXBP1*-RD related central nervous impairment/behavioral disturbance that would confound the scientific rigor or interpretation of results of the study<br>• History of intraventricular hemorrhage or structural brain deficit<br>• The presence of a clinical comorbidity deemed by the investigator to potentially confound the typical presentation of *SYNGAP1*-RD or *STXBP1*-RD<br>• Pregnant women or females of reproductive age who are found to be pregnant upon urine pregnancy testing |

We have enrolled 164 individuals with *STXBP1*-RD and 159 individuals with *SYNGAP1*-RD to date, ranging in age from 6 weeks to 68 years old. Clinical visits occurred over one or two days depending on the clinical site and/or the needs of the participant. We plan to follow a subset of about 200 individuals longitudinally, with an initial baseline visit, and follow-up assessments every 6 months.

### Developmental and parent-reported outcome measures

The selection of clinical and parent-reported outcome measures (PROMs) was completed in collaboration with the *STXBP1* Foundation and CURE SYNGAP1. An initial list of outcome measures was drafted to comprehensively assess relevant areas of impairment and functional limitation, as well as common medical complications associated with *STXBP1*-RD and *SYNGAP1*-RD. The complete list of assessments performed as part of the natural history study is outlined in **Table 2**, and includes scales and measures selected to capture all domains of development, including gross motor, fine motor, expressive and receptive communication, cognition, behavior, sleep, adaptive function, and symptoms of autism. These tools were chosen based on a landscape analysis of natural history study and clinical trial protocols for other similar DEEs (i.e., Dravet, Angelman), as well as literature review of *STXBP1*-RD, *SYNGAP1*-RD, and other DEEs with similar presentations. Site investigators who had clinical experience with *STXBP1*-RD and *SYNGAP1*-RD provided additional input. A series of meetings with the *STXBP1* Foundation and CURE SYNGAP1 reviewed the selected outcome measures and highlighted family-identified areas of concern. A final draft of the protocol was then approved by all parties. PROMs are administered through a third-party platform, RARE-X[25], starting from year two of the study once licensing agreements were finalized. We also obtained retrospective PROM data on individuals with *STXBP1*-RD and *SYNGAP1*-RD through Simon’s Searchlight for comparison with our prospective cohort.

**Table 2.**
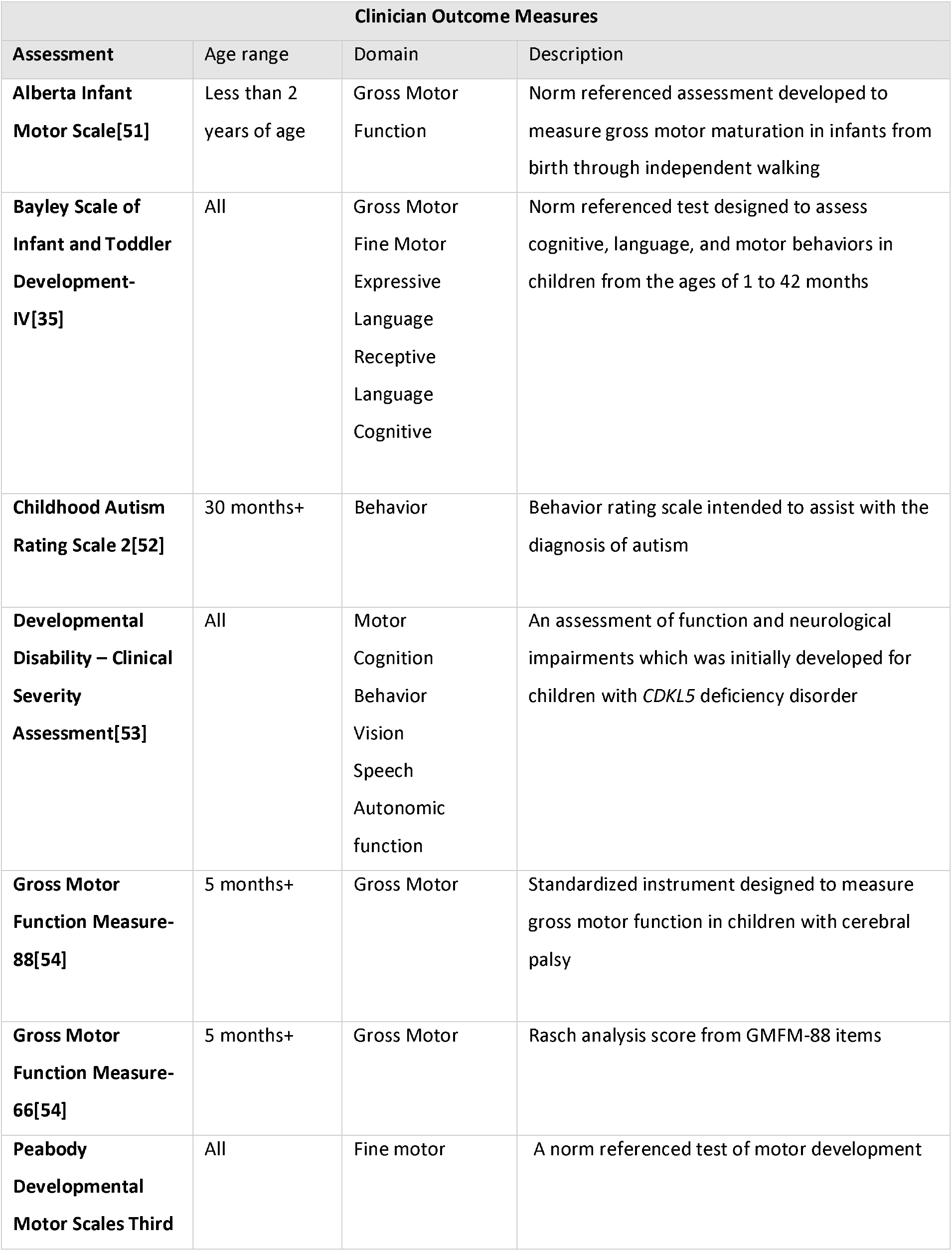
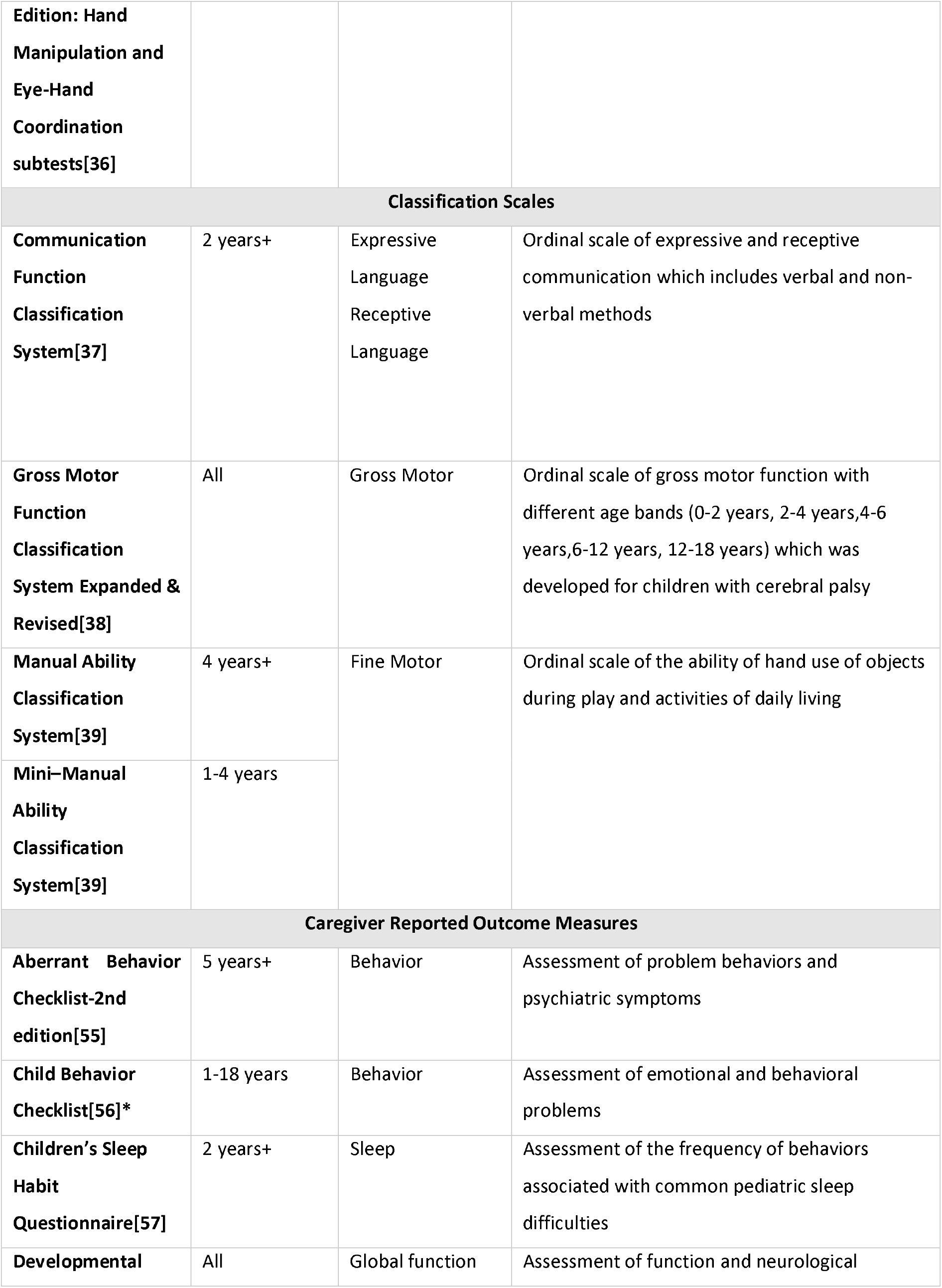
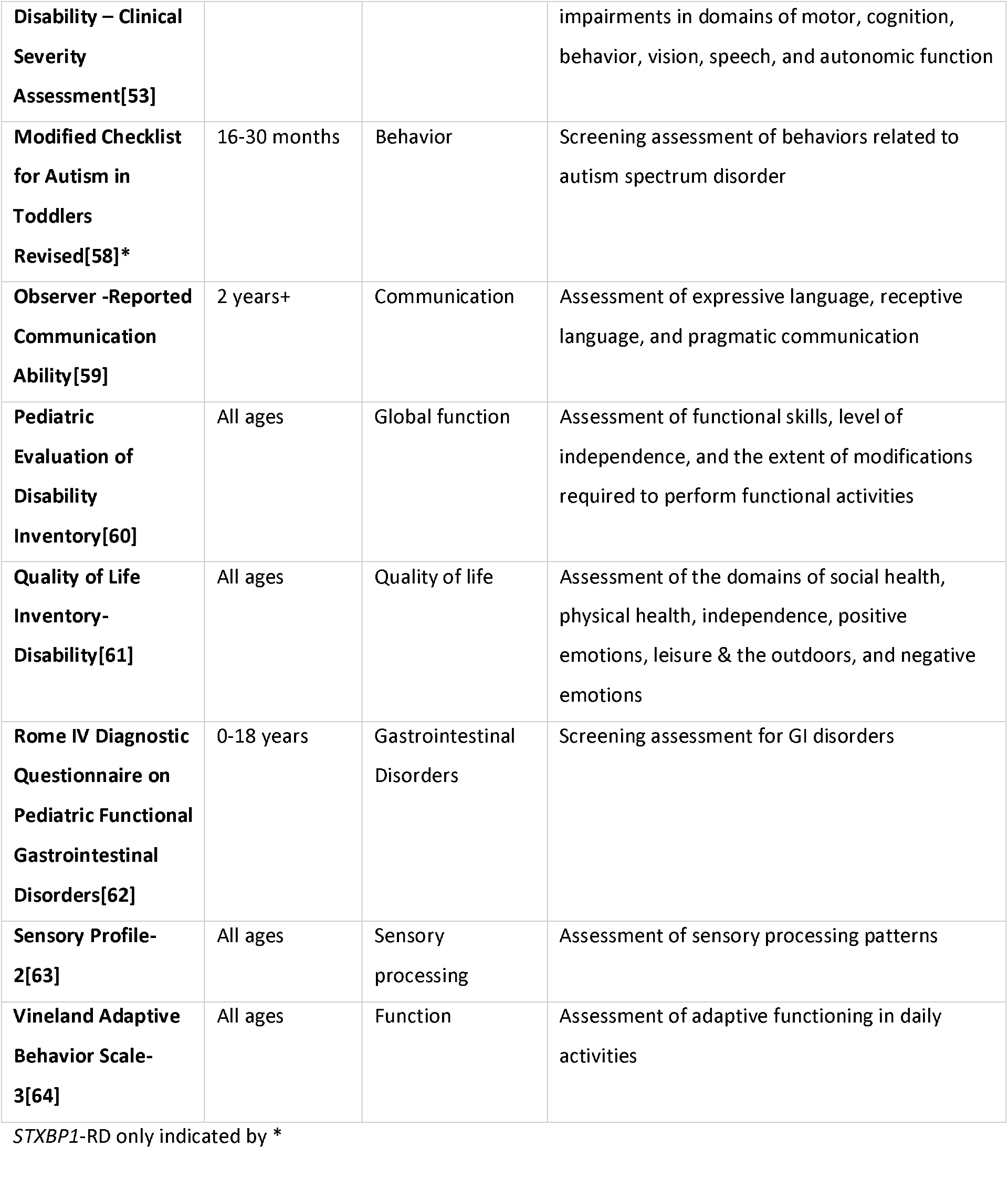
Outcome Measures.

#### Seizure history reconstruction

Seizure patterns for both disorders were documented using a standardized seizure reconstruction method previously described[5, 6, 26]. In brief, seizure frequencies and types were extracted from patient medical records and through detailed conversations with families during initial study visits. Following enrollment in the natural history study, seizure types and frequencies were documented and reconstructed on a month-by-month basis, with seizure type documented using Human Phenotype Ontology (HPO) terms and seizure frequencies converted to the Pediatric Epilepsy Learning Health System (PELHS) standardized scale[27, 28]. Of note, due to the high seizure frequencies often observed in *SYNGAP1*-RD, we developed an expanded PELHS scale, with two additional levels of 20-50 seizures per day and >50 seizures per day to better capture the variability in seizure frequency in this cohort. Future iterations of the protocol will include an app-based seizure diary.

#### Quantitative EEG

Resting state EEG data is acquired using a 128-channel Magstim EGI system and a cap-based electrode net, recorded at a sampling frequency of 1000 Hz. The testing is performed by trained clinical research staff and participants are seated comfortably in a quiet and dim room. Resting EEG data is recorded for 15-20 minutes. Records are annotated for eyes being open or closed, with any instances of sleep or excessive motion marked for eventual exclusion from the analysis. Visual (VEP) and auditory evoked potentials (AEP) are obtained during qEEG sessions. For VEPs, 400 trials of a reversing black and white checkerboard are presented. For AEPs, 520 trials of 500 Hz sinusoidal tones of 300 ms duration are presented at 60 dB using a free-field speaker. Distractions are minimized, and stimuli are presented at times when subjects are calm but attentive. All EEGs are downloaded from the EGI system and converted to European Data Format (EDF). To remove artifacts such as ocular movement, blinks, cardiac noise, and muscle artifact, an automated Independent Component Analysis (ICA) pipeline is applied[29, 30]. All scans have a harmonic 60Hz notch filter applied, followed by a 1-70 Hz bandpass filter. Data is analyzed in Python using a custom-built pipeline to extract the power spectra and power ratios across frequency bands.

#### Data Organization & Analysis

All data generated from the sites is entered into a 21 CFR Part 11–compliant REDCap database[31, 32], and data is exported and analyzed using RStudio. Figures were generated using the *ggplot2* package in R and the protocol figure (**Figure 1**) was created using BioRender.

## Preliminary Results

### Natural history studies in synaptic disorders are feasible with excellent participant retention

Since study initiation in July 2023, we have enrolled a total of 323 individuals into the combined natural history study. Of the 164 individuals with *STXBP1*-RD, 89 (54.2%) have returned for a second visit and 53 (32.3%) have had >2 assessments. 159 individuals with *SYNGAP1*-RD have been enrolled, with 73 (45.9%) returning for a second assessment and 32 (20.1%) completing >2 evaluations (**Figure 1**). We were able to recruit a large population despite the rarity of each disorder through collaboration with the *STXBP1* Foundation and CURE SYNGAP1. Recruitment has also been aided by the fact that the natural history assessments are conducted as part of a medical clinical visit, thus families receive concurrent clinical care, teaching, and recommendations. This incentivizes participation in the studies, while at the same time reducing burden. Importantly, for the longitudinal aim of the studies, retention has been better than expected, further highlighting community engagement.

### Retrospective seizure reconstruction generates consistent data without burden to families

*SYNGAP1*-RD and *STXBP1*-RD present notable challenges in seizure tracking, given that both disorders are notorious for the presence of difficult-to-count or uncountable seizures, such as eyelid myoclonia in *SYNGAP1-*RD and epileptic spasms in *STXBP1-*RD[1, 11]. In other prospective studies, collection of daily seizure counts through seizure diaries has been the traditional methodology, but significant challenges with completeness and burden on families have been noted[33, 34]. Therefore, we chose to track seizure data through a real-world data approach by reconstructing seizure types and frequencies through medical records, a method we have previously applied to other genetic epilepsies[4-6, 10, 26]. For preliminary analysis, seizure trajectories were reconstructed in a subset of 107 individuals with *STXBP1*-RD and 100 individuals with *SYNGAP1*-RD in monthly time bins totaling 902.7 patient years in *STXBP1*-RD and 950.3 patient years in *SYNGAP1*-RD (**Figure 2**).

**Figure 2.**
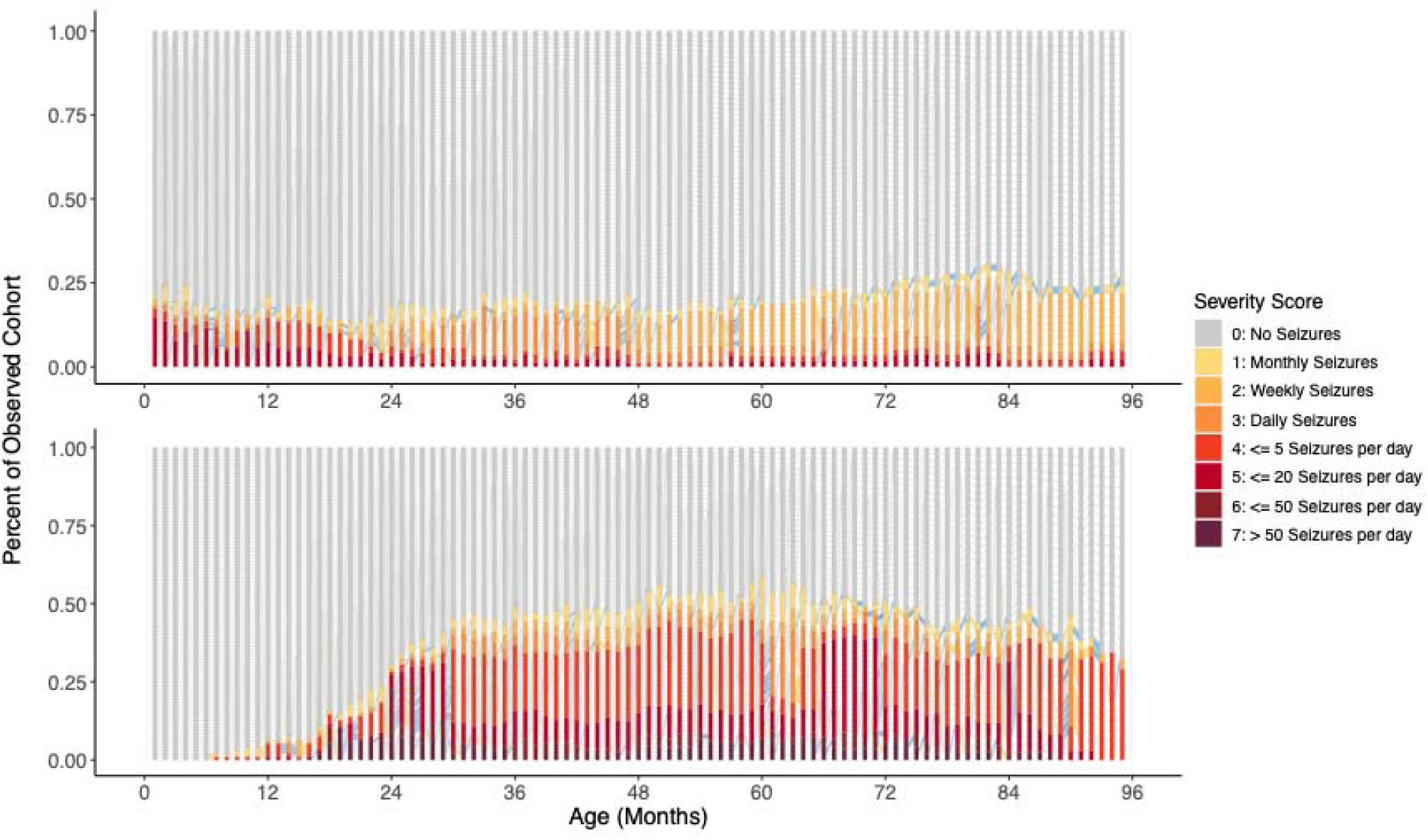
Retrospective seizure histories highlight the unique epilepsy trajectories in *STXBP1* and *SYNGAP1*. Seizure frequencies are recorded in monthly time bins from birth until the age at most recent assessment, and mapped onto the expanded PELHS frequency scale ranging from 0 (grey, no seizures) to 7 (burgundy, >50 seizures per day). Yellow and blue connecting lines indicate month-to-month worsening or improvement in seizure frequency, respectively. This analysis illustrates the relatively stable proportion of individuals with *STXBP1*-RD (***top***) with seizures over time, with some reduction in seizure frequency as individuals age. In contrast, individuals with *SYNGAP1*-RD (***bottom***) tend to have overall higher rates of active epilepsy and more frequent seizures, but with later onset– with seizures typically starting in the second or third year of life.

In *STXBP1*-RD, 73% (n= 78/107) of individuals experienced a seizure at any point in their lifetime, and ages of seizure onset ranged from day 0 of life through year 11.7, with median age of onset at 2.5 months. The most frequently experienced seizure types were focal motor seizures (n=61/107, 57.0%), epileptic spasms (n=27/107, 25.2%), and tonic seizures (n=26/107, 24.3%). At a cohort level, it was observed that within any window between 0 and 12 years of life, no more than 40% of the cohort had seizures at any one time point. In *SYNGAP1*-RD, 84% (n=84/100) of individuals had seizures at any point, with a median age of onset at 2.2 years. Most seizures were of generalized onset (78/100), including myoclonic seizures (52/100), eyelid myoclonia seizures (47/100), atonic seizures (45/100), and absence seizures (44/100) emerging as the most common types. Between the ages of 2 and 8 years, approximately 50% of the cohort was experiencing at least monthly seizures. Importantly, the frequency of seizures that could be considered countable in the context of a clinical trial was lower than the total seizure burden in both disorders. For *SYNGAP1*-RD, countable seizures were present in 20-45% of the cohort between the ages of 2-10 years and for *STXBP1*-RD, only about 25% of the cohort up until 10 years of age. Given countable seizures are not ubiquitously present across the cohort during target age ranges, exploration of other potential clinical trial outcomes measures is essential.

### Standardized clinical outcomes can be reliably obtained in individuals with synaptic disorders

Synaptic disorders present unique challenges in clinical outcome measure (COM) selection, as individuals most often present with severe global developmental delays and behavioral challenges that can make engaging in clinical assessments challenging. Creating a protocol that allows for completion in a clinic day and without significant floor or ceiling effects is paramount. We observed the appropriateness of several COMs, including the Bayley Scales of Infant and Toddler Development 4^th^ edition (Bayley-4), Gross Motor Function Measure-66 (GMFM-66)[35], and fine motor domains of the Peabody Developmental Motor Scales 3^rd^ Edition (Peabody-3)[36]. In addition, classification scales that were originally developed for children with cerebral palsy were used including Communication Function Classification System (CFCS)[37], Gross Motor Function Classification System Expanded & Revised (GMFCS)[38], Manual Ability Classification System (MACS)[39], and Mini–Manual Ability Classification System (miniMACS)[39]. As a preliminary analysis, we assessed completion as well as floor and ceiling effects at one study site where 89% (n = 87/106) of individuals with *STXBP1-* RD were able to complete the entire protocol, with 99% (n=105) of individuals being able to complete at least the GMFM-66. Similarly, 78% (n=80/116) of individuals with *SYNGAP1*-RD assessed at baseline were able to complete the entire protocol, and 99% (n=115) of individuals were able to complete at least the GMFM-66. Analysis of the GMFM-66 demonstrated distinct developmental patterns for both disorders, with individuals with *STXBP1*-RD on average showing lower scores of gross motor attainment than those with *SYNGAP1*-RD (**Figure 3**). Individuals unable to complete study measures were most often limited by behavior, somnolence, or travel scheduling constraints. Floor and ceiling effects, as defined by the number of individuals who achieved a minimum or maximum raw score, were negligible for *STXBP1*-RD (All Bayley-4 subtests: floor = 0%, ceiling = 0%; Peabody Hand Manipulation floor = 0%, ceiling = 0%; Peabody Eye Hand Coordination floor = 1% (n=1/87), ceiling = 0% GMFM-66 floor = 0%, ceiling = 0%) and for *SYNGAP1*-RD (Bayley Gross Motor floor = 0%, ceiling = 1% (n=1/87); Bayley Fine Motor floor = 0%, ceiling = 0%; Bayley Expressive Communication floor = 0%, ceiling = 6% (n=5/81;) Bayley Receptive Communication floor = 0%, ceiling = 0%; Bayley Cognitive floor = 0%, ceiling = 0%; Peabody Hand Manipulation floor = 0%, ceiling = 2% (n=2/94); Peabody Eye Hand Coordination floor = 0%, ceiling = 0% GMFM-66 floor = 0%, ceiling= 1% (n=1/115). Classification scales including the GMFCS, CFCS, and miniMACS/MACS and a novel severity scale (S-CSA) could be collected on all participants.

**Figure 3.**
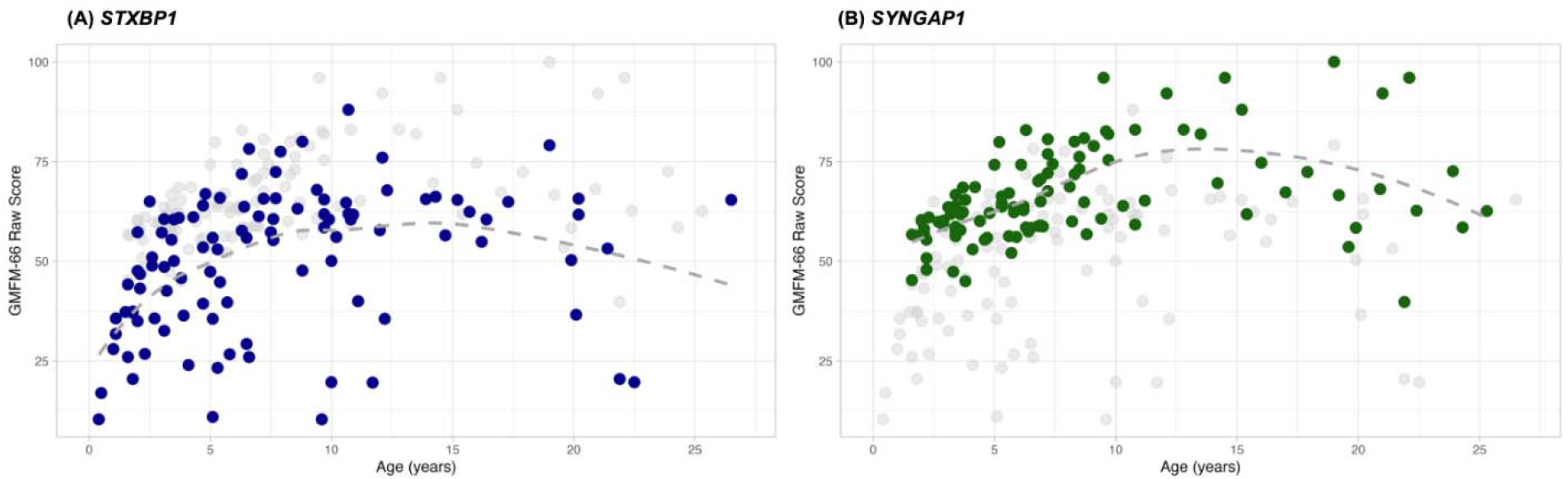
The GMFM-66 is a feasible gross motor assessment with little floor or ceiling effects. Initial GMFM-66 assessments for all individuals with *STXBP1*-RD (**A**, blue) and *SYNGAP1*-RD (**B**, green) are displayed across age. The grey dots in each plot represent the other condition to facilitate comparison. While individuals with *SYNGAP1*-RD tend to have higher gross motor function on the GMFM-66 when compared to *STXBP1*-RD, the variability in motor function of both disorders is appropriately captured by the measure, without significant floor or ceiling effects.

### Parent-reported outcomes can reveal disease burden not captured by clinical measures

Disease concept models (DCM) for both *STXBP1*-RD and *SYNGAP1*-RD have previously been completed, defining burden of disease from the perspective of parents with affected children[18, 19]. Combining this previous work with caregiver interviews and discussion with patient advocacy groups, domains of disease burden were defined and parent reported outcome measures (PROMs) were selected to ensure coverage of all disease domains defined through the DCMs. PROMs can offer insights into disease modalities that cannot be quantified in a clinical setting, including sleep, behavior, GI function, and quality of life impacts, and PROMs can offer data to complement COMs. To assess feasibility of selected PROMs, we combined existing data from the Simons Searchlight database for individuals with *STXBP1* (n=67) and *SYNGAP1* (n=39) with the emerging data from our natural history study collected using the RARE-X platform (**Figure 4**). Preliminary analysis of the Vineland-3 suggests that these scales are appropriate and feasible. While data is still insufficient to assess floor effects, our initial results suggest that these tests are feasible and can be completed by families. Further analysis is needed as our natural history study accumulates data on more participants. Completion rates of the measures by caregivers varied significantly, despite approaches such as in-person troubleshooting with trained staff and targeted discussion about the importance of completion with families.

**Figure 4.**
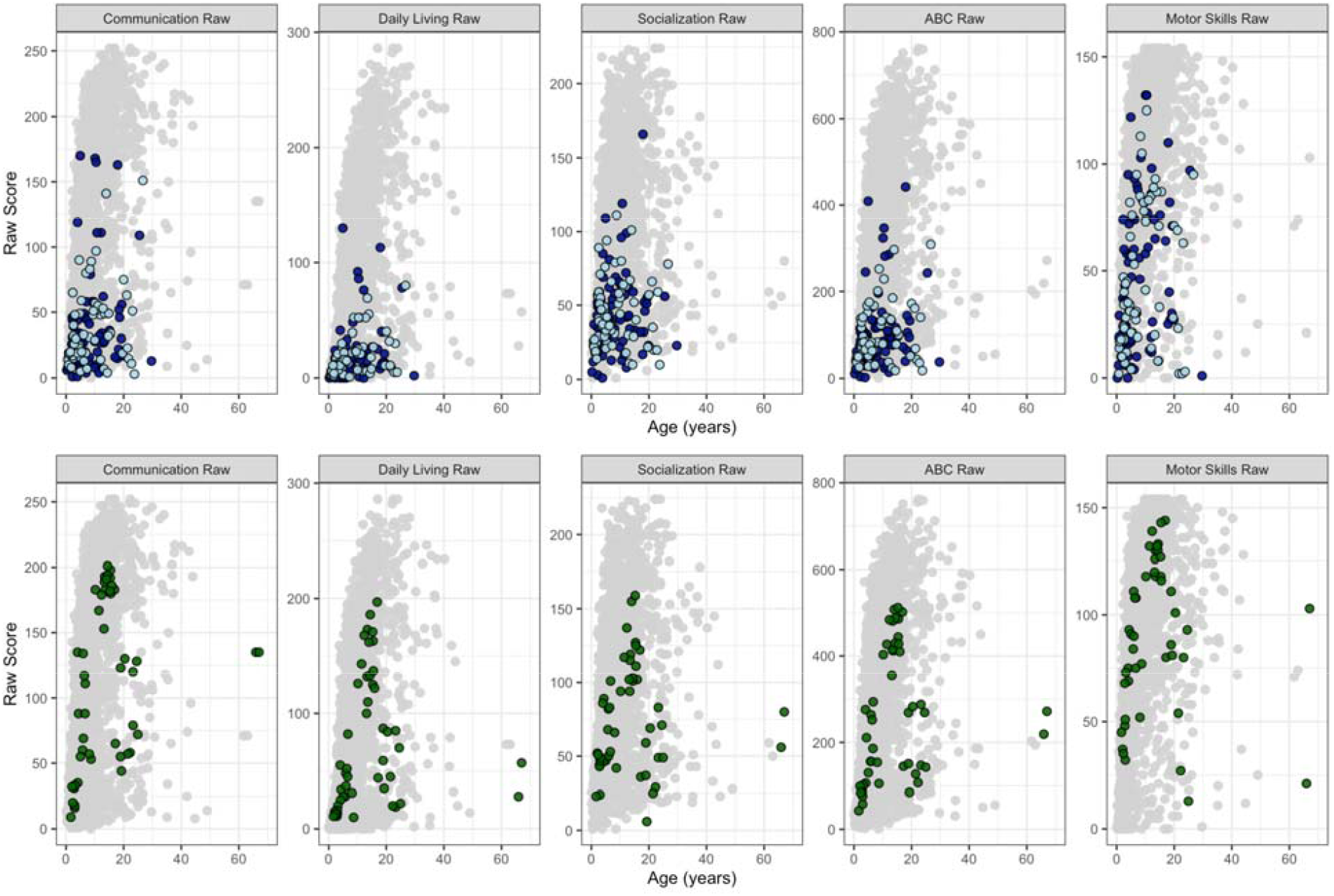
The Vineland-3 captures parent impressions of adaptive behavior across domains. Vineland-3 data from Simons Searchlight for *STXBP1*-RD (n=67, top, navy) and *SYNGAP1*-RD (n=39, bottom, green) was compared to all others in the Simons registry (1346 individuals across 107 disorders, grey). For *STXBP1*-RD, we also have preliminary data from RARE-X for our STARR NHS cohort displayed in light blue (n=45). The five domains are displayed separately, including communication, daily living, socialization, the ABC composite which is a sum of the prior 3 scores, and motor skills. Across domains, there is a broad range of adaptive functioning, with individuals with *SYNGAP1*-RD typically achieving higher performance than those with *STXBP1*-RD.

## Discussion

Here we present a protocol for a comprehensive natural history study for two common synaptic disorders, *STXBP1*-RD and *SYNGAP1*-RD, with detailed developmental, seizure, and behavioral assessments. Understanding the natural histories of both *STXBP1*-RD and *SYNGAP1*-RD is critical for future clinical trials which will attempt to improve the developmental trajectories, seizure frequency, medical co-morbidities, and behavioral challenges associated with these conditions. By carefully selecting the clinical assessments and PROMs for this NHS, we hope to learn detailed information regarding the appropriateness of various outcome measures for use in future clinical trials, as well as how feasible it is to collect a full battery of assessments in such complex patient populations.

Our study has three main findings. First, we demonstrate that the protocol, developed by an expert group of clinicians and patient advocates and informed by recent disease concept models, is feasible, with excellent completion rates across assessments and high participant retention rates. Second, we show through our initial data that existing outcome measures can be assessed in both *STXBP1*-RD and *SYNGAP1*-RD without significant floor or ceiling effects. The use of existing measures accelerates the process of obtaining natural history data and is currently favored by the FDA over the development of disease-specific measures[40]. Finally, our protocol is flexible, allowing for the future expansion to other DEEs, and the use of existing standardized measures facilitates data integration with other sources, comparisons to other disorders, as well as NHS expansion and future collaborations.

The collection of clinical outcome data for individuals with *STXBP1*-RD and *SYNGAP1*-RD presents several challenges. Many outcome measures, such as the Bayley-4[35], are norm-referenced to children of typical development, limiting the interpretation of test results for children with *STXBP1*-RD or *SYNGAP1*-RD, who may be older than the normative sample limit. As a result, the use of raw scores may be necessary, but these values may be limited by floor effects in children with significant motor and cognitive impairments[41]. Age equivalents are often reported in natural history studies[42], but these scores are imprecise and difficult to compare across individuals. As such, growth scale values may be preferred if available for any specific outcome measure[43]. It is also important that clinical outcome measures are sensitive to small changes, notable to families, that may result from an intervention[44]. Due to the difficulty and time required to develop new outcome measures, the FDA has suggested that natural history studies use or modify existing assessments if appropriate tools are available[40]. Here, we demonstrated the appropriateness of these existing tools in *STXBP1*-RD and *SYNGAP1*-RD, even when used out of age range, with no significant floor or ceiling effects noted. To determine if outcome measures should be modified to more specifically assess children with *STXBP1*-RD and *SYNGAP1*-RD, longitudinal assessments of a large sample of children are necessary, and this will continue to be evaluated during the longitudinal portion of the NHS. Since the course of development may be variable for children with *STXBP1*-RD and *SYNGAP1*-RD, the power to detect clinically meaningful and statistically significant differences between intervention groups necessitates careful consideration of outcome measures for a trial.

When designing the NHS, we decided to balance the competing interests of acquiring comprehensive data with minimizing the burden to families. This has resulted in several trade-offs, most critically, the use of seizure history reconstruction rather than seizure diaries, and the requirement for in-person assessments. In our prior work, we have demonstrated the feasibility of using retrospective medical records to reconstruct seizure trajectories in monthly time bins[5, 6]. Seizure types are coded by Human Phenotype Ontology (HPO) terms[45, 46] and frequencies are translated into the Pediatric Epilepsy Learning Health System (PELHS) seizure frequency score[27]. As demonstrated here, seizures in *STXBP1*-RD and *SYNGAP1*-RD are frequently “uncountable,” challenging accurate assessments of seizure burden. We do plan to pilot a digital seizure application commonly used in clinical trials to validate our retrospective assessment of seizure burden in both conditions and assess the feasibility of a seizure diary for use in future clinical trials. If counting seizures is not possible, trials may need to rely on EEG-based seizure counting or biomarkers as surrogate endpoints.

Another limitation of our study is the requirement for in-person assessments. Given the difficulty traveling with individuals with neurodevelopmental disorders, including the significant behavioral comorbidities associated with *SYNGAP1*-RD, it is possible that we are limiting the participation of those who reside far from study sites or have challenging behaviors or medical needs that preclude travel. Thus, we plan to pilot the Developmental Assessment of Young Children (DAYC)[47] in a subgroup of ProMMiS participants to validate its use compared to the Bayley-4. If successful, this would allow for remote participation in future clinical trials, alleviating family burden and allowing for assessment to occur in the individual’s natural home environment, minimizing stress and fatigue as confounders.

Another critical component to the design of clinical trials is the identification of robust biomarkers, as these enable precise patient stratification, improve trial efficiency, and increase the likelihood of detecting true therapeutic effects[48, 49]. Broadly, biomarkers are any measure that can separate individuals with a disorder from healthy controls (diagnostic), correlate with the severity of a disease (predictive), and show an improvement with disease-modifying therapy (responsive). Our current protocol represents an initial step toward realizing the full potential of biomarker discovery. While results were not discussed here, the NHS incorporates quantitative EEGs on all participants every 6 months, including auditory (AEP) and visual evoked potentials (VEP). Initial EEG analysis validates our prior findings that spectral features can serve as disease-specific biomarkers in *STXBP1*-RD and *SYNGAP1*-RD, with reduced alpha-theta ratios in the frontal and occipital regions, respectively, compared to healthy controls[50]. We are now evaluating the potential of event-related potentials (AEP and VEP) to serve as biomarkers of disease severity. Furthermore, we are exploring the diagnostic, predictive, and responsive potential of molecular biomarkers in serum, plasma and/or CSF from individuals with *STXBP1*-RD and *SYNGAP1*-RD.

## Conclusion

In summary, we have presented a comprehensive protocol for assessing natural history in *STXBP1*-RD and *SYNGAP1*-RD, including seizure trajectories, standardized developmental assessments, quantitative EEG, and parent-reported outcome measures assessing development, behavior, and sleep. We furthermore present initial data demonstrating that the protocol is feasible and that individual assessments capture the full range of outcomes without significant floor or ceiling effects. Therefore, this work represents a critical first step to make *STXBP1*-RD and *SYNGAP1*-RD clinical trial ready.

## Data Availability

Data produced in the present study are available upon reasonable request to the authors.

## Acknowledgments

This study was funded by the Center for Epilepsy and NeuroDevelopmental Disorders (ENDD) at Penn/CHOP, the NIH National Institute for Neurological Disorders and Stroke (R01 NS127830 and R01 NS131512 to IH and K23 NS140491 to JLM), the American Epilepsy Society (AES), Pediatric Epilepsy Research Foundations (PERF) & CURE SYNGAP1 through a Research Training Fellowship for Clinicians (JLM), and the American Academy of Neurology (AAN), AES, the Epilepsy Foundation, & the American Brain Foundation (ABF) through the Susan Spencer Award (JLM), the *STXBP1* Foundation through an individual grant (SMR) and the Career Ladder Education Program for Genetic Counselors grant from the Warren Alpert Foundation (SMR). Funding for investigators at Children’s Hospital Colorado, Texas Children’s Hospital, Rush University Medical Center, Stanford Medicine Children’s Health, and Weill-Cornell Medicine was provided by the *STXBP1* Foundation and for Children’s Hospital Colorado and Stanford Medicine Children’s Health by CURE SYNGAP1.

We would also like to thank patient advocates from CURE SYNGAP1 and the *STXBP1* Foundation for their contribution to protocol development and patient recruitment.

We appreciate the contributions of the additional site investigators at Baylor College of Medicine and Texas Children’s Hospital, including Alvina Zia, Ekaterina Sanchez-Romero, Elaine Seto, Danielle S. Takacs, Sruthi Thomas, Roberta Olivares, Natasha Feuerbach, Kristen S. Fisher, Mikael Guzman-Karlsson, Arden Wheeler, and Andres Jimenez-Gomez; at the Children’s Hospital Colorado, including Andrea Gerk, Megan Stringfellow, Brittany Gladfelter, Emily Wilson, Morgan Joliffe, Ann Reynolds, Kiara Hamlin, Julie Vanek, Kellie Sitarz, Kaitlyn Kennedy, and Dana Bennick; at Stanford Medicine Children’s Health, including Rayann Solidum, Sweta Patnaik, Amy Weisman, Sophia Magana, and Lauren Mattas; and at Weill-Cornell Medicine, including Dara Jones, Jennifer Cross, Carlianne Ward, Millie Stone, Natalie Wayland, and Aida Osis.

## Author Contributions

Jillian L. McKee – Conceptualization, Investigation, Formal Analysis, Funding Acquisition, Writing – Original Draft Preparation, Review & Editing

Sarah M. Ruggiero – Conceptualization, Investigation, Formal Analysis, Project Administration, Writing – Review & Editing

Kristin Cunningham – Investigation, Data Curation

JoeyLynn Coyne – Data Curation, Project Administration

Ian McSalley – Data Curation, Formal Analysis

Michael C. Kaufman – Data Curation, Formal Analysis

Bintou Bane – Data Curation, Project Administration, Writing – Review & Editing

Torrey Chisari – Data Curation, Project Administration

Jonathan Toib – Data Curation, Formal Analysis

Carlyn Glatts – Investigation, Data Curation

Sarah Tefft – Investigation, Data Curation

Julie Orlando – Investigation, Data Curation

Viveknarayanan Padmanabhan – Data Curation, Formal Analysis

Alexander K. Gonzalez – Data Curation, Formal Analysis

Alicia Harrison – Investigation, Data Curation

Charlene Woo – Investigation, Data Curation

Stephanie A. Zbikowski – Investigation, Data Curation

Rency Dhaduk –Data Curation, Project Administration

Johanna Mercurio – Data Curation, Project Administration

Macie McCarthy – Data Curation, Project Administration

Jan H. Magielski – Data Curation, Formal Analysis

Zachary Grinspan – Investigation, Supervision

Megan Abbott – Investigation, Supervision

Juliet Knowles – Investigation, Supervision

Hsiao-Tuan Chao – Investigation, Supervision

Katherine Xiong – Investigation, Supervision

Elizabeth Berry-Kravis – Investigation, Supervision

Sepideh Tabarestani – Investigation, Supervision

J. Michael Graglia – Funding Acquisition, Project Administration

Kathryn Helde – Funding Acquisition, Project Administration

Virgine McNamar – Funding Acquisition, Project Administration

Charlene Son Rigby – Funding Acquisition, Project Administration

James Goss – Funding Acquisition, Project Administration

Scott Demarest – Investigation, Supervision

Andrea Miele – Investigation, Supervision

Benjamin Prosser – Conceptualization, Funding Acquisition, Supervision

Michael Boland – Conceptualization, – Funding Acquisition, Supervision

Samuel Pierce – Conceptualization, Data Curation, Investigation, Writing – Original Draft Preparation

Ingo Helbig – Conceptualization, Investigation, Funding Acquisition, Writing – Review & Editing, Supervision

## Conflict of Interest/Ethical Publication Statement

Author Hsiao-Tuan Chao’s spouse has received support from Capsida Biotherapeutics, Inc.; author Ingo Helbig has received support from Capsida Biotherapeutics, Inc. The remaining authors have no conflicts of interest.

We confirm that we have read the Journal’s position on issues involved in ethical publication and affirm that this report is consistent with those guidelines.

